# Comparison of MRI sequences for optic nerve lesion detection in the follow-up of multiple sclerosis

**DOI:** 10.64898/2026.08.24.26361188

**Authors:** Máté Csomós, Magda Pribojszki, Brigitta Lóczi, Bence Bozsik, Nikoletta Szabó, Péter Faragó, András Király, Dániel Veréb, Eszter Tóth, Krisztián Kocsis, Krisztina Bencsik, László Vécsei, Zsigmond Tamás Kincses, Bálint Kincses

## Abstract

**Background:** Optic nerve involvement is common in multiple sclerosis (MS) and is now recognized as a key site for dissemination in space under the most recent revision of McDonald’s criteria. Reliable detection of optic nerve lesions is essential for diagnosis and monitoring, yet the optimal MRI sequence remains uncertain.

**Objective:** To compare the diagnostic performance of three MRI sequences—short tau inversion recovery (STIR), fat-suppressed FLAIR (fs-FLAIR), and double inversion recovery (DIR)—in detecting optic nerve lesions in MS patients.

**Methods:** Fifty-nine MS patients underwent MRI with STIR, fs-FLAIR, and DIR sequences and visual evoked potential (VEP) testing. Lesion detection was assessed independently for each sequence, and results were compared to structural and functional standards.

**Results:** No significant differences were found in lesion detection across the three sequences. All sequences showed similar sensitivity to structural and functional changes. The incremental benefit of adding orbita specific sequence to a whole-brain sequence was limited in the follow-up of MS.

**Conclusion:** In patients with established MS, whole-brain sequences (fs-FLAIR, DIR) perform comparably to dedicated orbital sequences (STIR) in detecting optic nerve lesions. This supports the feasibility of MRI protocols by omitting additional orbital sequences in routine follow-up, thereby reducing scan time and patient burden without compromising diagnostic sensitivity.

## Introduction

Regular follow-up of patients with multiple sclerosis (MS) is crucial for identifying disease progression and adapting treatment when necessary. Since MS activity may remain clinically silent, sensitive markers are needed to monitor involvement throughout the central nervous system. The optic nerve is one of the most commonly affected locations in MS (Graham & Klistorner, 2017), and its importance has recently been reinforced by its inclusion as a fifth anatomical site for dissemination in space in the updated diagnostic criteria (Montalban et al., 2025). Reliable assessment of optic nerve pathology is therefore increasingly relevant for both diagnosis and monitoring.

Techniques for reliably assessing the structural and functional integrity of the optic nerve are already available and are recommended for dedicated optic nerve evaluation, including optical coherence tomography (OCT), visual evoked potentials (VEP), and magnetic resonance imaging (MRI) (Barkhof et al., 2025; Montalban et al., 2025; Saidha et al., 2025) Although each method has distinct advantages and limitations (Saidha et al., 2025), the central role of MRI in MS diagnosis and follow-up (Wattjes et al., 2021) makes it particularly important to understand the sensitivity and specificity of different MRI sequences for detecting optic nerve involvement. Identifying the optimal sequence could shorten MRI protocols, thereby reducing costs and patient burden, especially in specialized MS centers where efficient and standardized imaging protocols are essential.

Recent expert recommendations support the use of optimized, dedicated MRI sequences for orbital imaging (Petzold et al., 2014; Wattjes et al., 2021). Visualization of the optic nerve is technically challenging because it is surrounded by orbital fat; therefore, fat suppression is required to improve lesion detection. Accordingly, fat-suppressed T2-weighted sequences or short tau inversion recovery (STIR) sequences are commonly recommended (Wattjes et al., 2021). However, both published reports and clinical experience suggest that other sequences, such as double inversion recovery (DIR), may also be useful for identifying optic nerve lesions (Healy et al., 2020a; Hodel et al., 2014). Although the optic nerve can be visualized and lesions can be detected with these approaches, it remains unclear which sequence is best suited for assessing optic nerve involvement, as their relative sensitivity has not been systematically compared.

To address this gap, we aimed to compare the diagnostic performance of three MRI sequences -STIR, fat-suppressed FLAIR (fs-FLAIR), and DIR - in detecting optic nerve lesions in patients with MS. By systematically evaluating structural findings against structural and functional evidence of optic nerve dysfunction, we aimed to determine which MRI sequence is most suitable for assessing optic nerve involvement.

## Methods

### Study overview

This cross-sectional study included participants from our MS centre (*Participants*). To evaluate the different MRI sequences in detecting optic nerve lesions, a blinded evaluator assessed each sequence independently (no knowledge of clinical status and history, *Assessment of MRI*). Identifying a gold standard measurement is challenging; therefore, we selected an approach to identify structural and functional standards as well. First, we used the radiologist’s unblinded MRI report from the routine clinical work-up as the structural reference standard (*structural standard*) (*MRI acquisition*; *Assessment of MRI*). Second, we used visual evoked potential (VEP) to characterize optic nerve integrity functionally (*functional standard*) (*Electrophysiological measurement*). We investigated the effect of sequences on the detection of optic lesions using these standards separately.

### Participants

Clinically stable relapsing-remitting MS patients (n=59) were recruited from our MS database (Bencsik et al., 2017) (Table 1). Patients were invited for an electrophysiological examination (*Electrophysiological measurement*), whose closest regular yearly check-up was within 5 weeks, and we used the closest regular clinical follow-up MRI scans for radiological evaluation (*Assessment of MRI*). The inclusion criteria were the absence of relapse within the six months preceding the VEP and MRI examinations and compliance with the American Clinical Neurophysiology Society guidelines for VEP assessment, including the absence of asymmetric pupil size (anisocoria). All patients were on disease-modifying therapy except for one. Full clinical data were available for all patients from our MS database (Bencsik et al., 2017), including a history of optic neuritis (ON). Healthy participants (HC; n=23) with no known neurological and psychiatric conditions were also recruited for electrophysiological examination to identify the threshold for normal values.

**Table 1.**
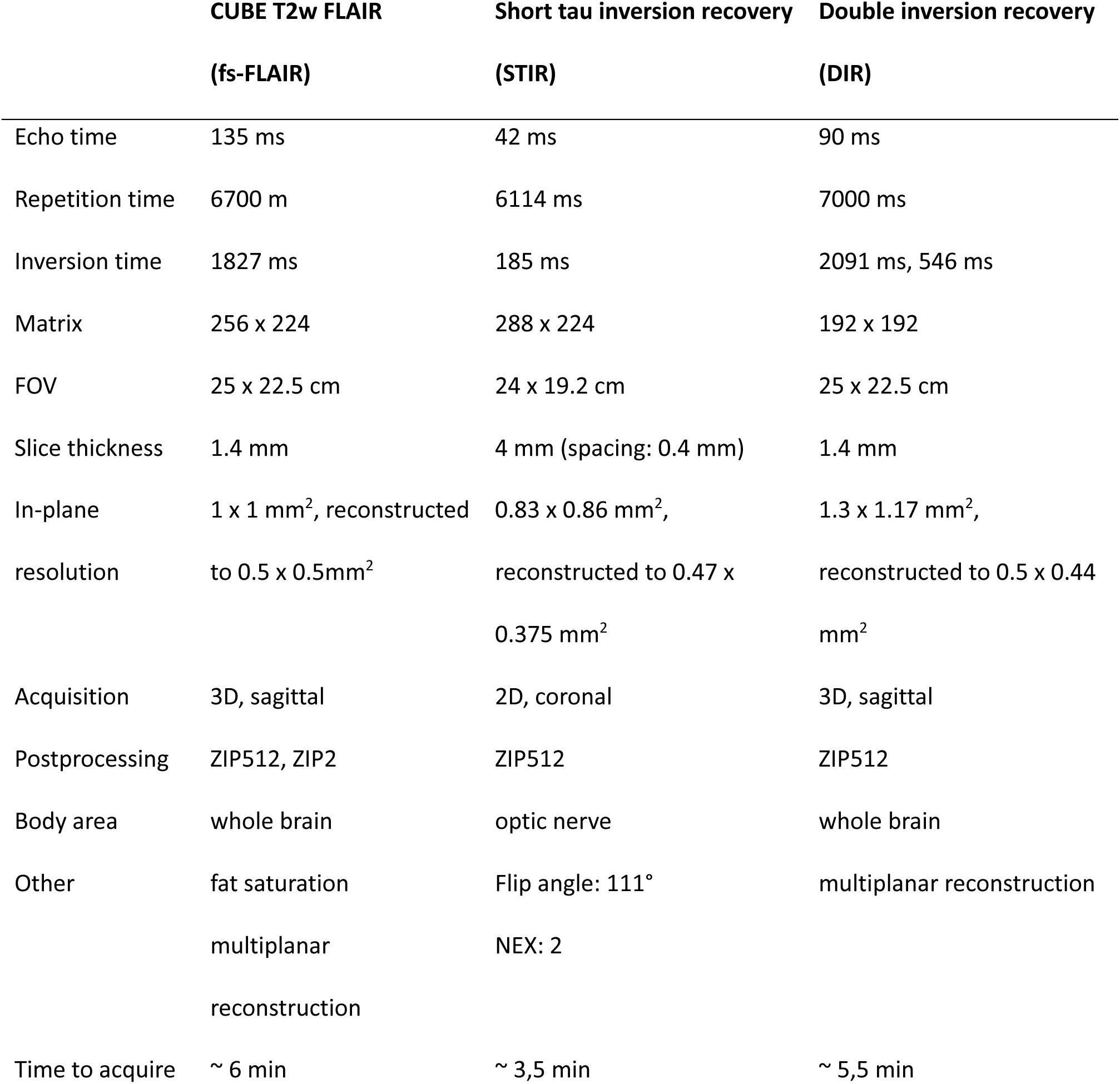
Acquisition parameters of the sequences used in this study

|  | **CUBE T2w FLAIR** | **Short tau inversion recovery** | **Double inversion recovery** |
| --- | --- | --- | --- |
|  | **(fs-FLAIR)** | **(STIR)** | **(DIR)** |
| Echo time | 135 ms | 42 ms | 90 ms |
| Repetition time | 6700 ms | 6114 ms | 7000 ms |
| Inversion time | 1827 ms | 185 ms | 2091 ms, 546 ms |
| Matrix | 256 x 224 | 288 x 224 | 192 x 192 |
| FOV | 25 x 22.5 cm | 24 x 19.2 cm | 25 x 22.5 cm |
| Slice thickness | 1.4 mm | 4 mm (spacing: 0.4 mm) | 1.4 mm |
| In-plane resolution | 1 x 1 mm<sup>2</sup>, reconstructed to 0.5 x 0.5mm<sup>2</sup> | 0.83 x 0.86 mm<sup>2</sup>, reconstructed to 0.47 x 0.375 mm<sup>2</sup> | 1.3 x 1.17 mm<sup>2</sup>, reconstructed to 0.5 x 0.44 mm<sup>2</sup> |
| Acquisition | 3D, sagittal | 2D, coronal | 3D, sagittal |
| Postprocessing | ZIP512, ZIP2 | ZIP512 | ZIP512 |
| Body area | whole brain | optic nerve | whole brain |
| Other | fat saturation<br>multiplanar reconstruction | Flip angle: 111°<br>NEX: 2 | multiplanar reconstruction |
| Time to acquire | ~ 6 min | ~ 3,5 min | ~ 5,5 min |

This study was carried out in accordance with the recommendations of the Medical Research Council National Scientific and Ethical Committee (ETT TUKEB) with written informed consent from all subjects. All subjects gave written informed consent in accordance with the Declaration of Helsinki. The protocol was approved by the National Institute of Pharmacy and Nutrition (000002/2016/OTIG).

### MRI acquisition

Magnetic resonance imaging was performed with a 3T GE Discovery 750w MR Scanner (GE Healthcare, Chalfont St. Giles, UK). MR images were acquired as part of routine clinical follow-up. The imaging protocol followed the recommendations available at the time and has been described in detail elsewhere (Kincses et al., 2018). The sequences used in the current study and their descriptors are summarised in Table 1. While the fs-FLAIR and DIR sequences provide whole-brain coverage, including the orbital region, STIR is a dedicated orbital imaging sequence.

### Assessment of MRI

A blinded evaluator (*MP,* radiologist with 5 years of experience) evaluated optic nerve involvement in each sequence (fsFLAIR, DIR, and STIR) separately. Importantly, the evaluator was blinded to all clinical and paraclinical information, and each sequence was assessed independently. The evaluation of FLAIR and DIR images was done after multiplanar reconstruction. A lesion in the optic nerve was defined as a hyperintensity that was observable along the prechiasmatic region (*Figure 1*). Additionally, a neuroradiologist (*ZTK*) evaluated all brain regions as part of the regular radiological work-up, that is he was aware of the participant’s clinical history, including prior episodes of optic neuritis, the results of paraclinical test (VEP, OCT), and had access to all available imaging sequences (fs-FLAIR, STIR, DIR), which may have influenced lesion detection and interpretation. This evaluation was used as a *structural standard* for optic lesions.

**Figure 1.**
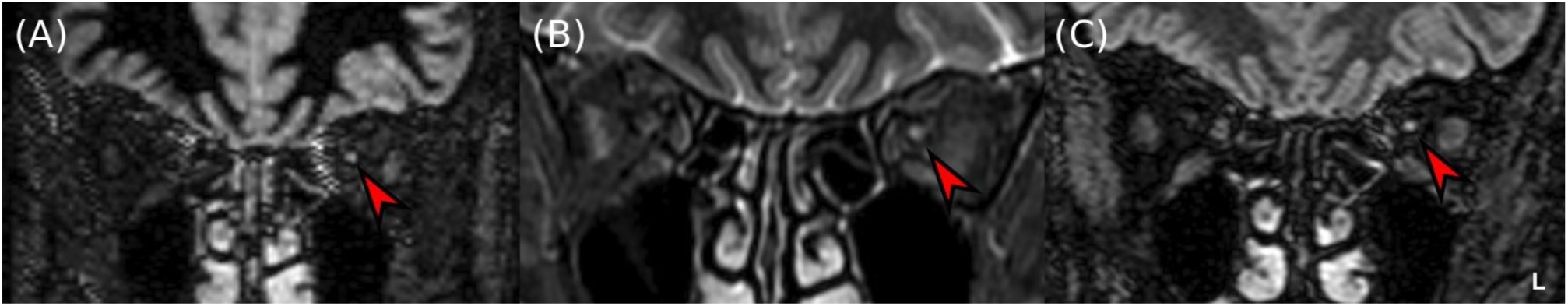
The three MRI sequences used in our analysis – fs-FLAIR (A); STIR (B); DIR (C)- in our analysis. The left optic nerve is hyperintense, which indicates an optic lesion. Note that the fs-FLAIR and DIR images are used after multiplanar reconstruction. The images are not in the same plane because of different slice thicknesses.

### Electrophysiological measurement

Visual evoked potential was used to assess functional integrity of the optic nerve (*functional standard*). Monocular pattern-reversal VEP was recorded (BrainAmp standard and actiCAP, Brain Products Inc.) following the guidelines of the American Clinical Neurophysiology Society (American Clinical Neurophysiology Society, 2006), using a black-and-white checkerboard stimulus (15’ and 60’ check sizes; small check data used for analysis) presented on a CRT screen. Recording electrodes were placed at Oz, O1, O2, and Fz (10/20 system), with the left mastoid as the reference and the vertex as the ground. P100 latency was determined offline (BrainVision Analyzer 2.0) from the Oz electrode (re-referenced to Fz) for each eye separately, yielding two values per participant. Normal limits derived from HC are used as thresholds for identifying functionally affected eyes (*Supplementary material: VEP classification and normal limits*). Additionally, recordings in which the P100 wave could not be reliably distinguished from noise were labelled as affected, and latency was not determined. Full stimulus and acquisition parameters are provided in the Supplementary material.

### Statistics

All the statistical tests were done in R Studio (version: 2025.9.0.387) (R Core Team, 2022). The code and data used for statistical analysis can be found in the Supplementary (Supplementary Code). Lesion detection performance of the different MRI sequences was investigated in relation to the standards (*structural and functional standards*). As all the data for the different assessments were in binary format (lesion present or not), diagnostic performance measures were primarily estimated directly from 2 × 2 contingency tables. We estimated different measures including specificity, sensitivity, positive and negative predictive values, and agreement (Cohen’s Kappa) for each sequence (fsFLAIR, STIR, DIR) separately. Cohen’s Kappa was interpreted as fair (0.21-0.4) or moderate (0.41-0.6) (Landis & Koch, 1977). Mixed-effects logistic regression was used for paired comparisons between MRI sequences while accounting for within-patient correlation. P-values were based on asymptotic normal approximations, and statistical significance was set at alpha = 0.05.

## Results

### Demographic

Demographic data of the participants are summarised in Table 2. The median expanded disability status score (EDSS) (Kurtzke, 1983) was 1. Patients received disease-modifying therapy except one due to breastfeeding. The mean disease duration was 90 months. Thirty-two patients with MS were found with a history of ON, with a total of 41 affected eyes (35% of the investigated optic nerves) and 25 with no history of ON (NON). Data on the history of ON were not available for two patients. The median months between the VEP and MRI measurements were 2 (range: 0-14). Participants had normal or corrected-to-normal visual acuity.

**Table 2.**
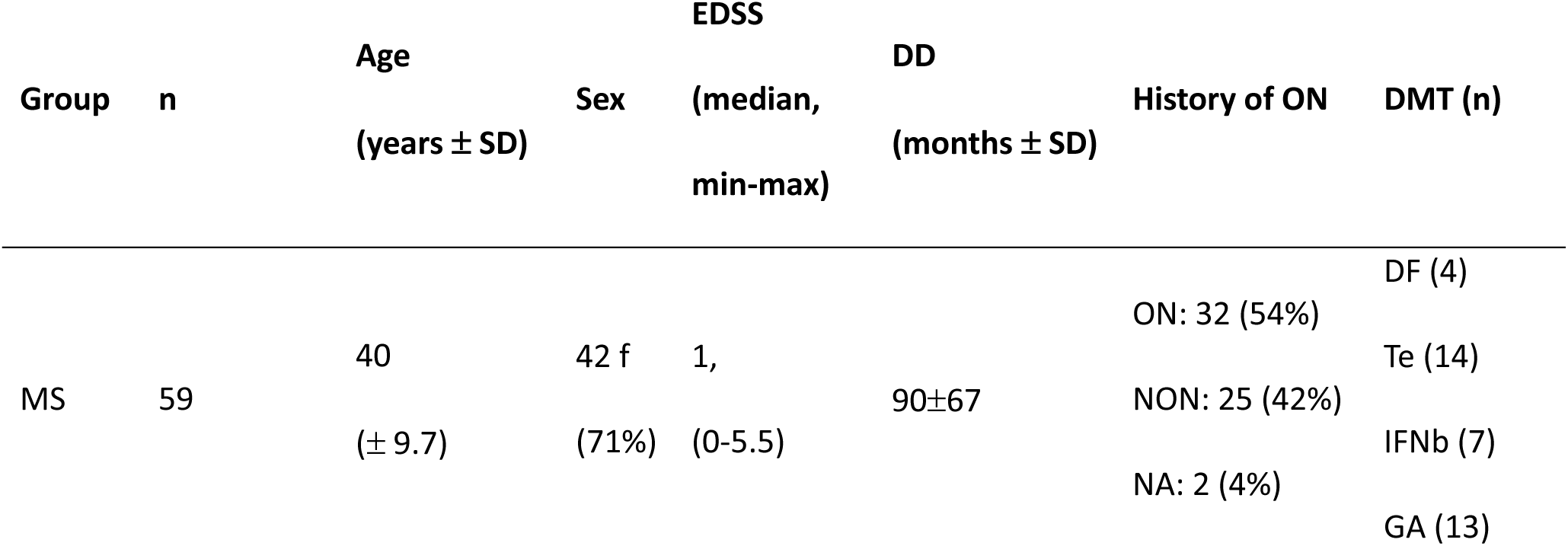
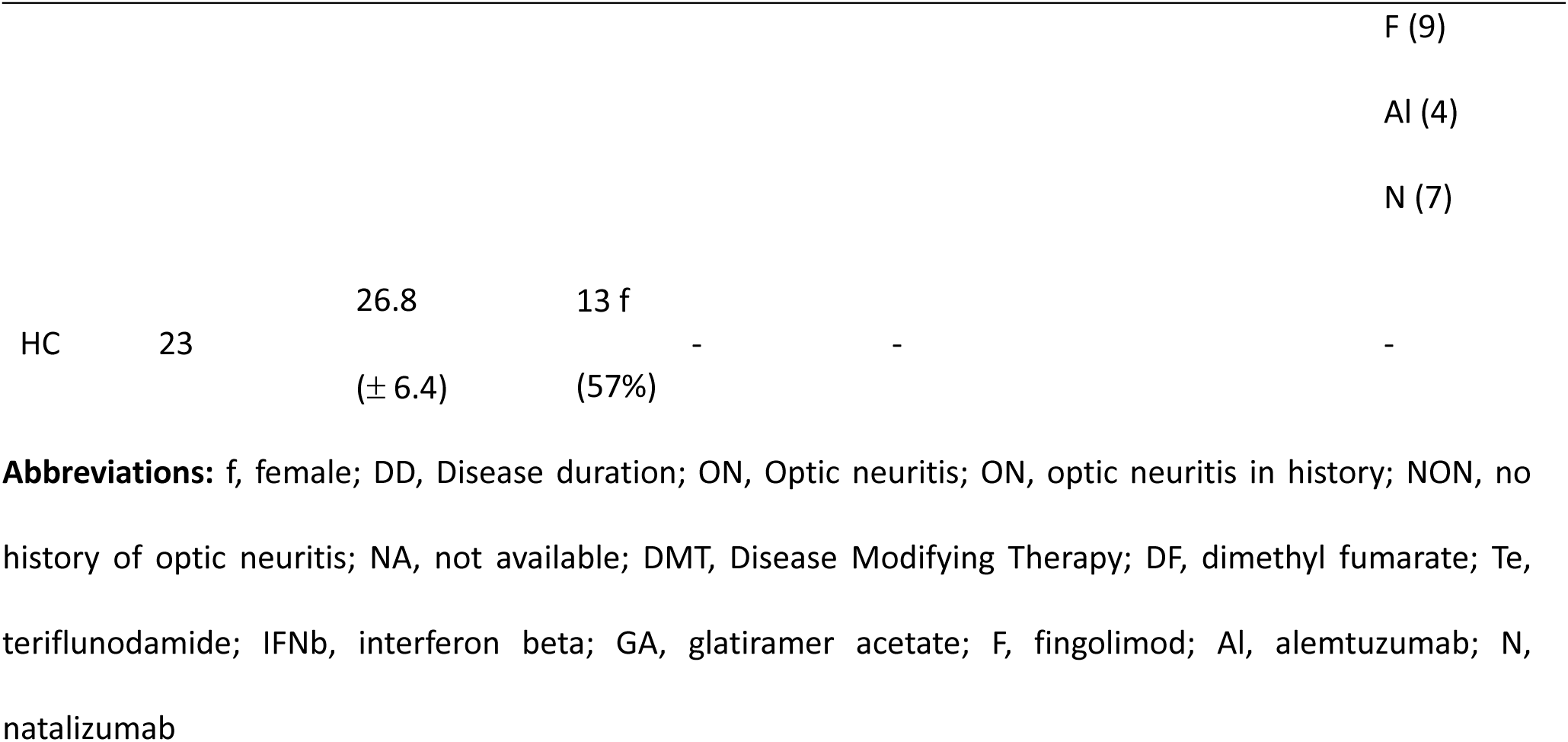
Demographic data of the cohort.

| Group | n | Age<br>(years ± SD) | Sex | EDSS<br>(median,<br>min-max) | DD<br>(months ± SD) | History of ON | DMT (n) |
| --- | --- | --- | --- | --- | --- | --- | --- |
| MS | 59 | 40<br>(± 9.7) | 42 f<br>(71%) | 1,<br>(0-5.5) | 90±67 | ON: 32 (54%)<br>NON: 25 (42%)<br>NA: 2 (4%) | DF (4)<br>Te (14)<br>IFNb (7)<br>GA (13) |

|  |  |  |  |  |  |  |
| --- | --- | --- | --- | --- | --- | --- |
|  |  |  |  |  |  | F (9) |
|  |  |  |  |  |  | AI (4) |
|  |  |  |  |  |  | N (7) |
| HC | 23 | 26.8<br>($\pm 6.4$) | 13 f<br>(57%) | - | - | - |

**Abbreviations:** f, female; DD, Disease duration; ON, Optic neuritis; ON, optic neuritis in history; NON, no history of optic neuritis; NA, not available; DMT, Disease Modifying Therapy; DF, dimethyl fumarate; Te, teriflunodamide; IFNb, interferon beta; GA, glatiramer acetate; F, fingolimod; AI, alemtuzumab; N, natalizumab

### The performance of different sequences in lesion detection

A representative example of a hyperintense optic nerve lesion detected on MRI is shown in Figure 1.

The relationship between the structural standard and lesion detection was assessed for each sequence. Overall, STIR showed the highest sensitivity, specificity, and predictive values, while fs- FLAIR and DIR performed similarly to one another and slightly lower than STIR across most measures. Moderate agreement was found for the STIR sequence (κ=0.49; 95% CI: 0.29-0.69; p<0.0001), but only fair agreement for the fs-FLAIR (κ=0.37; 95% CI: 0.17-0.57; p<0.0001) and DIR sequences (κ=0.40; 95% CI: 0.20-0.61; p<0.0001). The specificity, sensitivity, and predictive values are shown in Table 3 and Figure 2A. However, the overlapping confidence intervals suggest that the differences between sequences were not statistically robust, which was further supported by their non-significant difference estimated in the mixed-effects logistic regression (Figure 2B).

**Figure 2.**
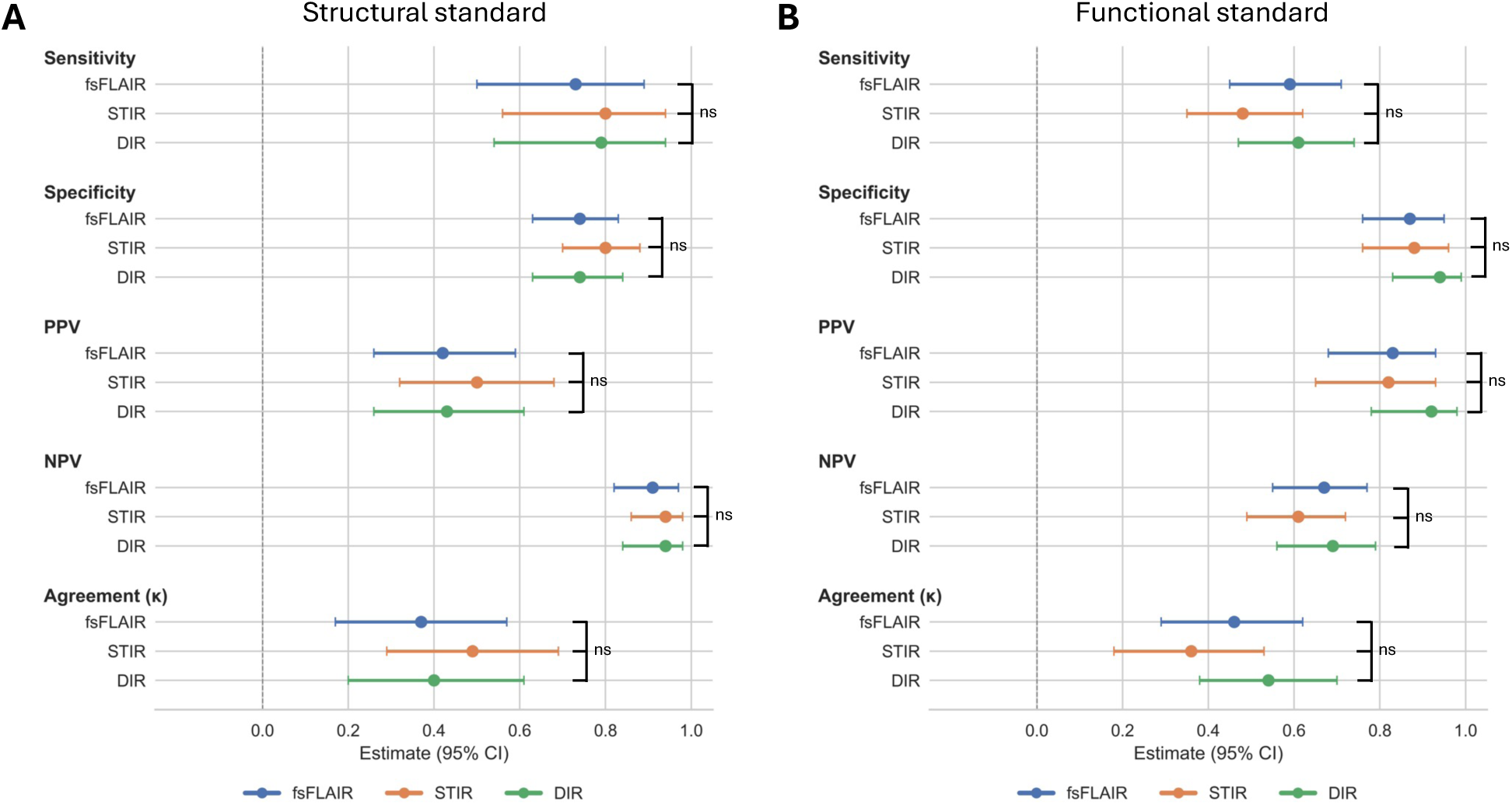
Diagnostic performance of MRI (rater MP) against (A) MRI and (B) VEP findings across the FLAIR, STIR, and DIR sequences. VEP served as the reference standard. Dots represent point estimates; Lines represent the 95% confidence intervals; Agreement refers to agreement with standard (Cohen’s κ). PPV, positive predictive value; NPV, negative predictive value; CI, confidence interval. ns – non significant difference.

**Table 3.**
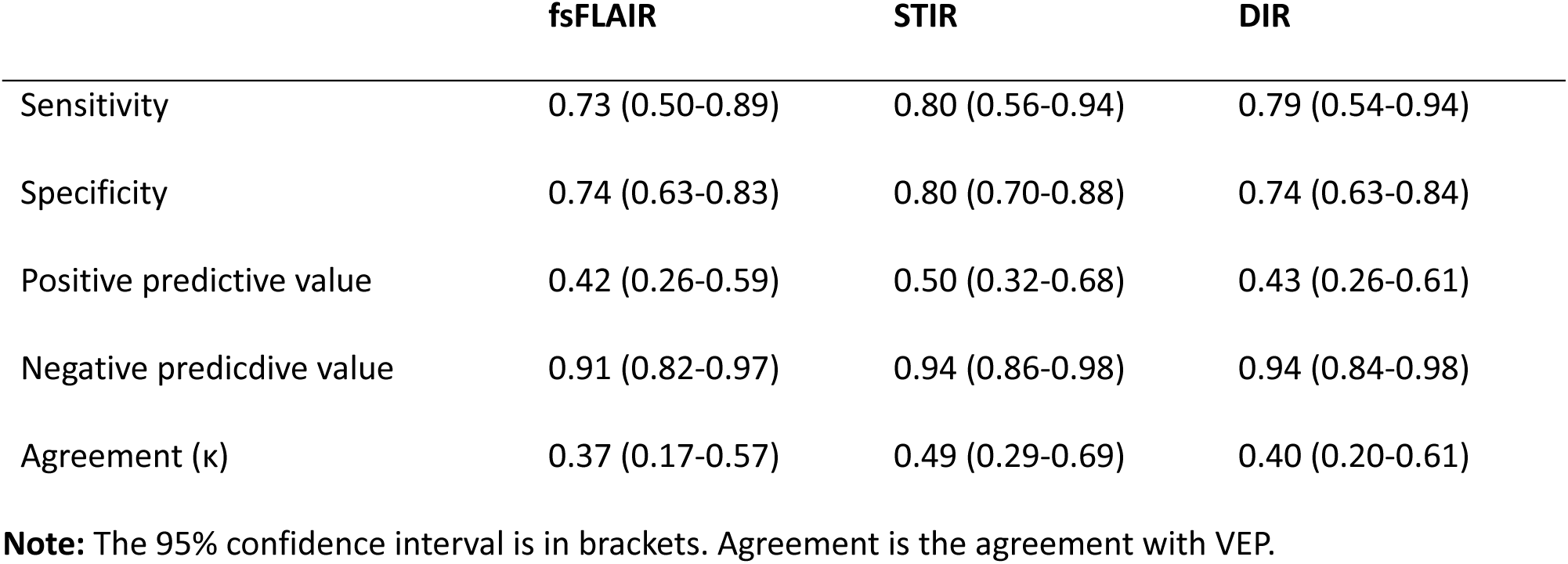
The association between MRI (blinded rater) and structural standard

The relationship between the functional standard and lesion detection was also assessed for each sequence. Overall, sensitivity was somewhat lower for STIR, while specificity was highest for DIR, though the remaining diagnostic measures were comparable across the three sequences. Moderate agreement was found for the DIR (κ=0.54; 95% CI: 0.38-0.7; p<0.0001) and fs-FLAIR (κ=0.46; 95% CI: 0.29-0.62; p<0.0001) sequences, but only fair agreement for the STIR sequence (κ=0.36; 95% CI: 0.18-0.53; p<0.0001). The specificity, sensitivity and predictive values are shown in Table 4 and Figure 2B. We found that the DIR sequence had the highest sensitivity, specificity, positive and negative predictive values among all the sequences. It has a high specificity (0.94 [95% CI: 0.83-0.99]) and a relatively low sensitivity (0.61 [95% CI: 0.47-0.74]); that is, it has a lower rate of false positives and a relatively higher rate of false negatives. However, the overlapping confidence intervals suggest that the differences between sequences were not statistically robust, which was further supported by their non-significant difference estimated in the mixed-effects logistic regression (Figure 2B).

**Table 4.** The association between MRI (*rater MP*) and functional standard

|  | fsFLAIR | STIR | DIR |
| --- | --- | --- | --- |
| Sensitivity | 0.59 (0.45-0.71) | 0.48 (0.35-0.62) | 0.61 (0.47-0.74) |
| Specificity | 0.87 (0.76-0.95) | 0.88 (0.76-0.96) | 0.94 (0.83-0.99) |
| Positive predictive value | 0.83 (0.68-0.93) | 0.82 (0.65-0.93) | 0.92 (0.78-0.98) |
| Negative predictive value | 0.67 (0.55-0.77) | 0.61 (0.49-0.72) | 0.69 (0.56-0.79) |
| Agreement ($\kappa$) | 0.46 (0.29-0.62) | 0.36 (0.18-0.53) | 0.54 (0.38-0.7) |

**Note:** VEP findings were used as a reference to evaluate the performance of MRI findings. The 95% confidence interval is in brackets. Agreement is the agreement with VEP.

## Discussion

In the present study, we evaluated the performance of three MRI sequences – fat-supressed FLAIR, STIR, and DIR - in the detection of optic nerve lesions in patients with multiple sclerosis (MS). Our results indicated that no single sequence consistently outperformed the others across all assessed metrics. This suggests that whole brain high-resolution sequences, which include the orbital region as well, provide comparable performance to a dedicated orbital sequence in the detection of optic nerve lesions in the follow-up of MS.

The inclusion of the optic nerve as a distinct anatomical site in the revised McDonald criteria underscores its critical role in the diagnosis of MS (Montalban et al., 2025). As one of the most commonly affected pathways in MS, the visual system is involved in a substantial proportion of patients - optical neuritis (ON) presents as the initial clinical manifestation in approximately 25% of cases (A. T. Toosy et al., 2014). Moreover, individuals with isolated ON have a 50% risk of developing clinically definite MS within 15 years, a risk that rises to 75% if white matter lesions are detected on baseline MRI (The Optic Neuritis Study Group, 2008). Throughout the disease course, more than two-thirds of MS patients experience at least one episode of ON (A. T. Toosy et al., 2014). Given the high prevalence and clinical significance of optic nerve involvement, the development of robust, reliable, and sensitive imaging methods for detecting and monitoring optic nerve pathology is essential for early diagnosis, prognosis prediction, and treatment monitoring.

Different MRI sequences are available for reliably detecting optic nerve lesions, each with distinct advantages for visualising inflammatory and demyelinating pathology. In the context of acute optic neuritis (ON), lesions are typically associated with contrast-enhancing signal abnormalities (Kupersmith et al., 2002), reflecting active inflammation. However, growing concerns regarding the cumulative use of gadolinium-based contrast agents in longitudinal MS monitoring have prompted a shift toward non-contrast imaging sequences. The MAGNIMS guideline recommends fat-suppressed T2 or STIR for routine assessment (Wattjes et al., 2021). While technical parameters (e.g.: slice thickness) have been explored (Healy et al., 2020b), direct comparisons across sequences remain limited. DIR imaging has demonstrated superior sensitivity for acute ON lesions compared to combined STIR and FLAIR (Hodel et al., 2014) and it is proven to be effective in detecting subclinical optic nerve pathology (Hadhoum et al., 2016; Riederer et al., 2019; Sartoretti et al., 2017). In accordance with these findings, all the sequences showed in our study a good sensitivity and specificity in optic nerve lesion detection. However, we found no statistically significant differences in lesion detection performance across fs-FLAIR, STIR, and DIR sequences. This suggests that all three sequences demonstrated comparable sensitivity for identifying optic nerve pathology in our clinical cohort, and the added value of a dedicated orbital sequence may be limited in the follow-up of MS. These findings have meaningful implications for clinical MRI practice. Notably, the STIR sequence is limited to the orbital region, whereas fs-FLAIR and DIR are acquired as part of a whole-brain protocol, covering both the optic nerves and the entire brain parenchyma. This distinction translates into a significant time difference: acquiring a dedicated STIR sequence adds approximately three minutes to the scan duration. Shortening the scan protocol by omitting the additional orbital sequence - particularly in follow-up assessments - could reduce total scan time by approximately 10% and decrease patient burden, without compromising lesion detection. Given that whole-brain sequences with adequate resolution perform comparably to dedicated orbital imaging in identifying structural changes linked to visual dysfunction, they may suffice for routine monitoring of disease progression in MS. However, high-resolution orbital sequences remain valuable in the initial diagnostic workup, especially for differential diagnosis - particularly when distinguishing MS from conditions such as neuromyelitis optica spectrum disorders. Thus, a protocol using high-resolution whole-brain imaging may be sufficient for monitoring, reserving dedicated orbital sequences for diagnostic uncertainty or acute presentations.

Our study focused on MRI-based assessment, and we used an unblinded radiological evaluation of all MRI images as the structural reference standard. This may introduce potential interrater variability in lesion detection as two different raters (standard and blinded for the comparison of sequences) were included in the study. To mitigate this limitation, we incorporated VEP as a more objective functional measure - despite its known constraints, including sensitivity to non-optic lesions and a variable dissociation between structural and functional integrity (Backner et al., 2019; Behbehani et al., 2017; Davies et al., 1998; Graham & Klistorner, 2017; Masi et al., 2016; Thurtell et al., 2009; Walt et al., 2015). Nevertheless, the convergence of structural (MRI) and functional (VEP) data across sequences supports the conclusion that no single MRI sequence consistently outperforms the others. Additionally, optic nerve assessment can be complemented by tools such as OCT and VEP, yet the optimal approach - whether a single modality suffices or a combined strategy is necessary - remains unresolved (A. Toosy et al., 2026). Although MRI, OCT, and VEP provide complementary insights, their relative contributions - particularly in the context of sequence selection and clinical phase - remain unclear. This underscores the need for future studies to systematically compare multimodal outcomes in both diagnostic and follow-up settings, using blinded, standardized assessments to further clarify the optimal imaging strategy.

## Conclusion

Optic nerve involvement is common in multiple sclerosis - even subclinically - and its monitoring is essential for assessing disease progression. MRI remains a key tool for evaluation, though technical challenges due to the optic nerve’s complex anatomical environment in the orbit necessitate optimized imaging strategies. Our findings show no significant difference in lesion detection among fs-FLAIR, STIR, and DIR sequences, suggesting that whole-brain sequences alone may suffice for routine assessment. This supports protocols by omitting additional sequences, thereby reducing scan time and participant burden without compromising sensitivity. These results highlight the feasibility of using high-resolution whole-brain imaging with appropriate sequences for effective, efficient monitoring of optic nerve pathology in MS.

## Data Availability

All data produced in the present study are available upon reasonable request to the authors

## Acknowledgements

The authors thank András Kincses for his methodological assistance with the MRI sequences.

