## Supplementary for "Comparison of MRI sequences for optic nerve lesion detection in the follow-up of multiple sclerosis"

### Methods

#### Electrophysiological measurement

All VEP measurements were performed with the same parameters across participants according to the guidelines of the American Clinical Neurophysiology Society with an EEG system (BrainAmp standard and actiCAP, Brain Products Inc). Monocular VEP was recorded during black and white checkerboard pattern reversal stimulation. Stimuli were presented on a CRT screen, distanced 0.9 m from the participants, with two check sizes of 15 and 60 minutes of arc. The total field size was 21º in the horizontal plane and 15º in the vertical plane. Data from the small check condition was used for the analysis. Luminance and Michelson contrasts were 58 cd/m2 and 63%, respectively. 200 reversals were recorded in total, at 2 reversals/s. Recording electrodes were placed on the Oz, O1, O2 and Fz according to the international 10/20 system. Reference and ground were on the left mastoid and on the vertex, respectively. After the recording, offline post-processing was done to evaluate P100 latency from the Oz electrode with BrainVision analyzer 2.0 (Brain Products Inc). First, Oz was re-referenced to Fz, and a 1-40 Hz bandpass filter was used; then segmentation and baseline correction were performed. Lastly, an average of the segments and semi-automatic peak detection were performed to estimate the latency of the P100 wave. P100 latency of the left and right eyes was estimated separately for the small check size condition, which resulted in two values per subject. All P100 waves were checked, and the automatic peak detection was corrected manually in case of error. Moreover, recordings on which the typical VEP wave could not be separated from the noise were labelled as affected, and P100 latency was not determined

#### VEP classification and normal limits

The normal distribution of the HC P100 latencies was examined with the Shapiro-Wilk test. Using HC data, the similarity of the left and right eyes was investigated with the Bland-Altman diagram. The range of agreement was defined as mean bias ±2SD, and the grand average was calculated if there was a significant similarity between the two eyes. Mean and standard deviation (SD) of P100 and interocular latencies were estimated in HCs. These estimates were used to determine normal limits as mean±2SD. P100 and interocular latencies of the MS participants were compared to these limits and classified as prolonged or normal and used as a *functional standard*.

#### P100 latency differences across groups

To investigate P100 latency differences across groups, data from each eye were considered separately (two values per participant), and a mixed model analysis was employed, in which the ON history of the eyes was used as a fixed factor and subjects as a random factor. When a significant main effect was detected, pairwise comparisons were conducted using Tukey's post hoc test.

### Results

#### Visually Evoked Potentials

To establish the normal range for our setup, the following steps were taken. The Bland-Altman test showed that the P100 latency was similar between the left and right eyes in healthy control (HC) participants, so a grand mean was computed. The HC mean latency was 101.2 ms (SD: 5.1 ms). The upper limit was then set as the mean plus two standard deviations, which is 111 ms. Similarly, the interocular latency difference limit was determined, resulting in an upper threshold of 7 ms. Patient latencies and interocular differences were compared to these limits, with higher latencies indicating a positive diagnosis on this *functional standard*. An example comparison between a patient with MS and a HC can be seen in Fig. S1.

Figure S1 demonstrates a prolonged P100 wave in a patient with multiple sclerosis compared with a healthy control.

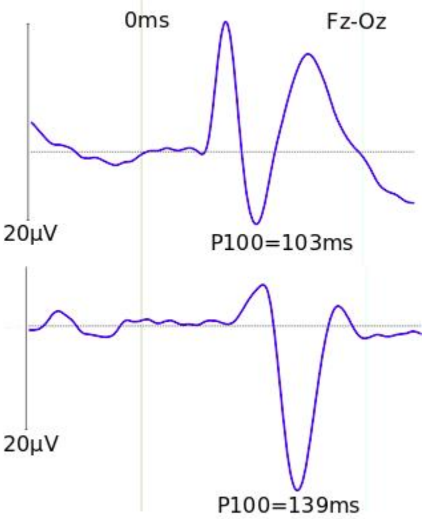


**Figure S1** Two VEP registrations from the re-registered and pre-processed (to Fz, see Methods) Oz electrode, for demonstration purposes. The upper registration is from a male healthy participant, while the lower is from a female multiple sclerosis patient. The grey sidebar on the left represents 20µV. The vertical light grey line shows the zero time point. A prolonged P100 wave could be observed in an MS patient with a peak at 139ms. Note that the MS participant's registration has a relatively large amplitude, which might partly be due to the sex difference.

In patients with multiple sclerosis, VEP results were positive in 49 eyes and negative in 59 eyes based on latency criteria. Nine eyes of seven participants had abnormal waveforms and could not be distinguished from noise. In these cases, the VEP diagnosis was considered positive. One recording (one participant‘s one eye) was not available because of hardware-related issues (Figure S2).

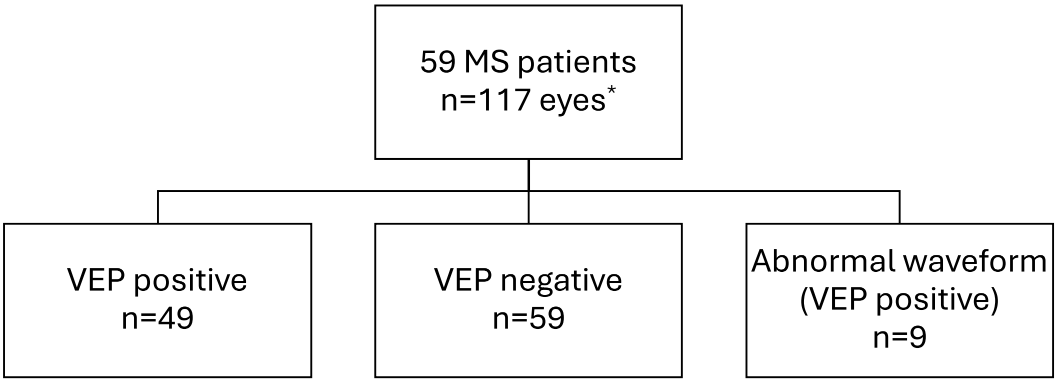


**Figure S2** VEP findings in the eyes of patients with multiple sclerosis. *One recording of one eye was contaminated.

Latency of eyes with ON history, eyes without ON history, and HCs' eyes were compared (Figure S3). Eyes with different ON history had prolonged latency (F(2,107.6)=16.3; p<0.0001), moreover, post-hoc tests revealed significant differences between HC and all other groups and between affected eyes and the other two groups (Tukey post hoc test: HC vs. ON (101ms vs. 117ms): t(99.4)=5.5; p<0.001; HC vs. NON (101ms vs. 110ms): t(80.3)=3.2.; p<0.01; ON vs. NON (110ms vs. 117ms): t(107)=3.9; p<0.001).

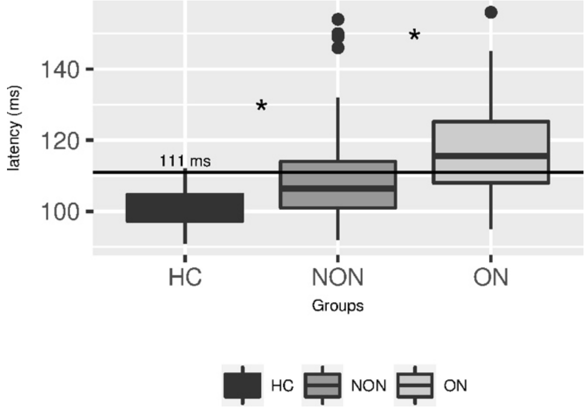


**Figure S3** Latency of P100 wave in visual-evoked potential. The multiple sclerosis group had a prolonged latency compared to healthy controls. Moreover, an eye with a history of optic neuritis had the longest latency. Asterisks stand for significant differences (p<0.05) between groups. The upper threshold (111ms), based on the HC data, is illustrated. HC - healthy controls (n=23); NON - no optic neuritis in history (n=25); ON - optic neuritis in history (n=32).

#### Association between structural (MRI) and functional (VEP) standards

The association between structural (MRI) and functional (VEP) standards were also investigated with a 2x2 contingency table. There was fair agreement between the two methods: κ=0.24 (95% CI: 0.06-0.42, p<0.005). The sensitivity and specificity were 0.32 (95% CI: 0.2-0.46) and 0.92 (95% CI: 0.82-0.98), respectively. The positive and negative predictive values were 0.82 (95% CI: 0.6-0.95) and 0.56 (95% CI: 0.45-0.67), respectively (Table S1).

**Table S1** 2x2 contingency table of structural (MRI) and functional (VEP) standards

| MRI optic lesion \ VEP finding | positive | negative |
|---|---|---|
| positive | 18 | 4 |
| negative | 38 | 49 |
